# DEFINE: A Phase IIa Randomised Controlled Trial to Evaluate Repurposed Treatments for COVID-19

**DOI:** 10.1101/2021.05.20.21257513

**Authors:** E Gaughan, T Quinn, A Bruce, J Antonelli, V Young, J Mair, A Akram, N Hirani, O Koch, C Mackintosh, J Norrie, J Dear, K Dhaliwal

## Abstract

**Introduction:** COVID-19 (Coronavirus Disease 2019) is a new viral-induced pneumonia caused by infection with a novel coronavirus, SARS CoV2 (Severe Acute Respiratory Syndrome Coronavirus 2). At present there are few proven effective treatments. This early phase experimental medicine protocol describes an overarching and adaptive trial designed to provide safety, pharmacokinetic (PK)/ pharmacodynamic (PD) information and exploratory biological surrogates of efficacy, which may support further development and deployment of candidate therapies in larger scale trials of COVID-19 positive patients.

**Methods and analysis:** DEFINE is an ongoing exploratory multicentre platform, open label, randomised study. COVID-19 positive patients will be recruited from the following cohorts; a) community cases b) hospitalised patients with new changes on a chest x-ray (CXR) or a computed tomography (CT) scan or requiring supplemental oxygen and c) hospitalised patients requiring assisted ventilation. Participants may be recruited from all three of these cohorts, depending on the experimental therapy, its route of administration and mechanism of action.

The primary statistical analyses are concerned with the safety of candidate agents as add-on therapy to standard of care in patients with COVID-19.

**Safety will be assessed using:**

- Haematological and biochemical safety laboratory investigations.
- Physical examination
- Vital signs (blood pressure/heart rate/temperature and respiratory rate)
- Daily electrocardiogram (ECG) readings
- Adverse events

The analysis population will consist of (i) all patients randomised to a treatment arm who receive any dose of the study drug and (ii) all patients randomised to the control arm who would also have been eligible to receive a study drug.

Secondary analysis will assess the following variables during treatment period 1) the response of key exploratory biomarkers 2) change in WHO ordinal scale and NEWS2 score 3) oxygen requirements 4) viral load 5) duration of hospital stay 6) PK/PD and 7) changes in key coagulation pathways.

**Ethics and dissemination:** The DEFINE trial platform and its initial two treatment and standard of care arms have received full ethical approval from Scotland A REC (20/SS/0066), the MHRA (EudraCT 2020-002230-32) and NHS Lothian and NHS Greater Glasgow and Clyde.

The results of each study arm will be published as soon as the treatment arm has finished recruitment, data input is complete and any outstanding patient safety follow-ups have been completed. Depending on the results of these or future arms, data will be shared with larger clinical trial networks, including RECOVERY, and to other partners for rapid roll out in larger patient cohorts.

**Registration details:** The DEFINE protocol has been registered on ISRCTN (https://www.isrctn.com/) and Clinicaltrials.gov(https://www.clinicaltrials.gov/).

ClinicalTrials.gov Identifier: NCT04473053

ISRCTN Identifier: ISRCTN14212905

**Strengths and limitations of this study:**

- The trial is as flexible as possible to ensure a broad range of patients can be recruited and candidate therapies can be added or removed as evidence emerges.
- The team are collecting real world data of medications at an early stage of their use in COVID-19 across the full spectrum of disease; allowing the administration of different treatment formulations (inhaled vs oral vs intravenous).
- The simultaneous collection of clinical outcomes as well as exploratory endpoints including clinical biomarkers, flow cytometry, PK/PD and thromboelastography allows further characterisation and elucidation of the temporal immuno-inflammatory cascade in COVID-19 to inform on future therapy selection.
- This is a Phase 1b/IIa platform study and thus the primary end point is clinical safety therefore our anticipated numbers will be too small to allow for definitive data on efficacy.
- DEFINE is an experimental medicine platform, currently restricted to three clinical sites and so the generation of data will be slower than that of larger platforms with access to a greater number of patients.

## Introduction

COVID-19 (Coronavirus Disease 2019) is a new viral-induced pneumonia caused by infection with a novel coronavirus, SARS CoV2 (Severe Acute Respiratory Syndrome Coronavirus 2). This highly contagious, zoonotic infection was first identified in Wuhan, China in late 2019^1^ and its emergence has led to a global pandemic with a significant impact on health, society and the economy.

The majority of those infected with SARS CoV2 have asymptomatic or mild infection however 10-20% will require admission to hospital with hypoxaemic respiratory failure requiring oxygen therapy and possibly ventilatory support^2^. Risk factors for severity include increased age, obesity and past medical history such as diabetes and hypertension^3^. Genetic predisposition is now known to play a role^4^ and much of the lung damage is driven by a surge of inflammatory mediators. Mortality rate amongst hospital in-patients is felt to be as high as 20%^5^. By May 2021 over 3 million individuals worldwide have died of COVID-19. To date there are few proven treatments with significant impact upon mortality and despite multiple successful vaccination developments it is expected that the virus will become endemic in the population, with significant levels of disease activity for years to come.

DEFINE is a phase Ib/IIa experimental medicine trial. This protocol describes an overarching and adaptive trial designed to provide safety, pharmacokinetic (PK)/ pharmacodynamic (PD) information and exploratory biological surrogates of efficacy which may support further development and deployment of candidate therapies in larger scale trials of COVID-19 positive patients receiving normal standard of care.

Given the spectrum of clinical disease, community based infected patients or hospitalised patients can be included. Products requiring parenteral administration will only be investigated in hospitalised patients. Patients will be divided into cohorts as described below. Participants may be recruited from all three of the study cohorts, depending on the experimental therapy, its route of administration and mechanism of action.

Candidate therapies can be added to the protocol and previous candidates removed from further investigation as evidence emerges. The trial will be monitored by an independent Data Monitoring Committee (DMC) to ensure patient safety.

Each candidate cohort will include a small cohort of patients randomised to candidate therapy or existing standard of care management dependent on disease stage at entry. Each treatment arm will recruit up to 20-30 patients to provide safety data, and PK-PD profile of potential therapeutic agents against COVID-19.

As COVID-19 follows a variable clinical path in individual patients, the protocol is designed to enable inclusion of patients across the disease stages. The trial is intended to provide mechanistic data from patients receiving standard of care therapy and from patients treated with the therapy candidates. The study will enable delivery of pharmacokinetic information and effects of standard of care and candidate agents on surrogate biomarkers of the disease process and the specific drug target.

## Methods and analysis

### Study design

DEFINE is an exploratory multicentre platform, open label, randomised study. This experimental medicine platform trial encompasses early phase studies, and the identification of major safety signals is the primary objective. Each candidate therapy will include a small cohort of patients randomised to the candidate therapy or existing standard of care management dependent on disease stage at entry. Randomisation will be computer generated at a ratio of 1:1 across arms.

#### Cohorts

Patients with confirmed SARS CoV2 infection with relevant COVID-19 symptoms or signs will be recruited into this trial. As SARS CoV2 has a range of clinical manifestations, representatives of three target patient cohorts will be included in this trial. Participants may be recruited from one or more of these cohorts, depending on the experimental therapy under investigation. While the study will approach patients with suspected COVID-19 and conduct screening assessments, only patients who are confirmed SARS-CoV-2 positive will be randomised.

|  |  |
| --- | --- |
| **Cohort 1A** | Community (primary care) patients with confirmed COVID-19 |
| **Cohort 1B** | Community (primary care) patients with confirmed COVID-19 with new changes on CXR or CT scan compatible with COVID-19 and deemed 'high-risk' of hospitalisation or death* |
| **Cohort 2A** | Hospitalised confirmed COVID positive patients with new changes on CXR or new changes on CT compatible with COVID-19 but not requiring supplemental oxygen, |
| **Cohort 2B** | Hospitalised confirmed COVID positive patients with: new changes on CXR or new changes on CT compatible with COVID-19 and requiring supplemental oxygen , |
| **Cohort 3** | Hospitalised patients with confirmed COVID-19 requiring assisted ventilation (including non-invasive and mechanical ventilation) |

\* High risk is defined as over 50 years of age with comorbidities.

#### Eligibility criteria

Eligible participants were hospital inpatients over the age of 16 with confirmed SARS CoV-2 infection within 14 days of a positive test. Exclusion criteria includes pregnancy, lactation, and the inability to reliably take or tolerate modes of treatment delivery, or if the patient was receiving anticoagulation, antiplatelet therapies or potassium sparing diuretics which could not reasonably be withheld. Patients were also excluded if they had a current or recent history of severe, uncontrolled cardiac disease (NYHA class IV), diabetes mellitus, renal impairment (eGFR <30 or ongoing dialysis) or hepatic impairment (ALT >5x upper limit of normal), anaemia with a Hb <80, PLT <50, hyponatraemia with a Na <120 or K+ >5.0. Co-enrolment with a Clinical Trial of an Investigational Medicinal Product (CTIMP) will not be permitted.

#### Concomitant treatment

Any drug required for the normal clinical care of these patients will be permitted, although as treatment assets are added to the DEFINE platform interactions with candidate therapies will be reviewed on a case by case basis.

### Objectives and Endpoints

#### Primary

The primary outcome is to evaluate the safety and tolerability of candidate agents as add-on therapy to SoC in patients with COVID-19. Safety will be assessed using daily:

- Haematological and biochemical safety laboratory investigations
  - Full blood count, urea, creatinine, sodium, potassium, alanine transaminase, aspartate transaminase, alkaline phosphatase, coagulation screen (including APPT, PT, INR, fibrinogen), glucose
- Physical examination (if clinically relevant)
- Vital signs
  - blood pressure, heart rate, respiratory rate, oxygen saturations, oxygen requirement, temperature
- Electrocardiogram (ECG) readings
- Adverse events

#### Secondary

- To explore the PK/PD or appropriate surrogate of bioavailability of the proposed trial treatments in COVID-19 patients.
- Assess the response of key exploratory biomarkers during treatment period. Evaluate the change from baseline values for key exploratory biomarkers of target engagement for each treatment.
- To evaluate the improvement or deterioration of patients in each treatment arm using the WHO ordinal scale and NEWS2 score.
- To evaluate the number of oxygen-free days (e.g. duration (days) of oxygen use and oxygen-free days).
- To evaluate ventilator-free days and incidence and duration of any form of new ventilation use. This will be measured in duration (days) of ventilation and ventilation-free days. Incidence of any form of new ventilation use and duration (days) of new ventilation use.
- Change in the ratio of the oxygen saturation to fraction of inspired oxygen concentration (SpO2/FiO2). SpO2/FiO2 will be measured daily from first dose to Day 15, hospital discharge, or death.
- To evaluate SARS-CoV-2 viral load. Qualitative and quantitative polymerase chain reaction (PCR) determination of severe acute respiratory syndrome coronavirus 2 (SARS-CoV-2) in saliva samples while hospitalised on Days 1, 3, 5, 8, 11, 15 and oropharyngeal/nasal swab on the same days if tolerated.
- To evaluate time to discharge and duration to discharge following treatment.
- To evaluate the use of renal dialysis or haemofiltration for each treatment arm.

#### Follow up

All discharged patients will undergo follow up to assess for adverse events at 30 days, 60 days and 90 days.

### Data collection and analysis

The biomarkers variables will be mostly continuous measures and the treatment effect estimated via a linear model which will adjust for baseline covariates highly correlated with the primary outcome, including possibly the baseline measurement of the primary outcome biomarker.

The study will not be powered for subgroup analyses and these will be exploratory, on a limited number of subgroups pre-specified in the study protocol.

Due to the small sample sizes there will be no formal adjustment for missing data, and the primary analysis set – appropriate for early phase proof of signal studies – could be a suitably defined per-protocol set (e.g. those that were compliant with their randomised medications).

Safety data will be analysed on the as-treated data-set (anyone who initiated on randomised treatment) and will be presented descriptively.

The independent DMC will review accumulating data, unblinded to the randomised groups. Their first and foremost responsibility will be the safety of the participants, and the committee may terminate the study at any time on the grounds of safety.

### Data management

The Principal Investigator is responsible for the quality of the data recorded at each Investigator Site.

All Investigators and study site staff involved with this study must comply with the requirements of the appropriate data protection legislation (including where applicable the General Data Protection Regulation with regard to the collection, storage, processing and disclosure of personal information). Access to personal information will be restricted to individuals from the research team treating the participants, representatives of the sponsor(s) and representatives of regulatory authorities.

Computers used to collate the data will have limited access measures via user names and passwords.

Published results will not contain any personal data that could allow identification of individual participants.

## Ethics and dissemination

For the DEFINE trial, a number of ethical and safety considerations have been addressed.

1. The DEFINE trial recruits adults with severe COVID-19 symptoms who are often incapacitated. The selection and enrolment of adults with incapacity will take place within the legal framework described in Adults with Incapacity (Scotland) Act 2000 and Medicines for Human Use (Clinical Trials) Regulations, 2004. During the pandemic, relatives were not allowed to visit the hospital so the majority of consent for adults with incapacity will be undertaken over the phone. This is a well-practiced method of obtaining consent and will be undertaken following all necessary guidance and with a set of instructions to ensure the personal legal representative has all the appropriate information and safeguarding in place. In rare cases if no appropriate personal legal representative can be found, and it is believed to be in the best interests of the patient, consent from a professional legal representative may be sought. This is a person not connected with the trial who is either the doctor primarily responsible for the patient’s medical treatment, or is a person nominated by the relevant health care provider.
2. Patients may not be able to physically sign the consent forms themselves as this could increase potential transmission of COVID-19. The researcher will be able to in these instances sign the consent form on behalf of the patient once verbal consent is taken.
3. Trial treatments have been extensively tested pre-clinically and in patients prior to DEFINE. All necessary safety information regarding the treatments is available to the research team and will be referred to in case of any serious safety event. All risks relating to the treatments will be carefully explained to both patients and their representatives.
4. Inclusion and exclusion criteria will carefully considered to avoid exposing patients to undue risk.
5. All appropriate approvals will be in place prior to the start of recruitment.

Scientific publications and the sharing of clinical data generated as part of this trial is crucial to better understanding COVID-19 and developing new treatments. As such, the results of each study arm will be published as soon as the treatment arm has finished recruitment, data has been cleaned and any outstanding patient safety follow-ups completed.

## Data Availability

Scientific publications and the sharing of clinical data generated as part of this trial is crucial to better understanding COVID-19 and developing new treatments. As such, the results of each study arm will be published as soon as the treatment arm has finished recruitment, data has been cleaned and any outstanding patient safety follow- ups completed.
External datasets and supplementary online material will be avaialble.

## Authors’ contributions

All of the authors have contributed to the initial study protocol and the protocol paper.

